# Who Can Get Functional Neurological Disorder and How Do They Get It? Pathophysiological Insights from Epidemiological Data

**DOI:** 10.64898/2026.05.01.26352255

**Authors:** David D.G. Palmer, Mark J. Edwards, Jason B. Mattingley

## Abstract

**Background and Objectives:** Functional neurological disorder (FND) is one of the most common causes of neurological symptoms and disability, but much remains unknown about its pathophysiology. In both clinical conversations and research publications, clinicians and researchers imply a variety of models for onset of the condition with respect to both the process culminating in its onset, and the distribution of susceptibility to the condition across the population. Here we used population-level data as evidence to arbitrate between these generative models of the condition.

**Methods:** We identified six hazard distributions corresponding to different pathophysiological processes, and four distributions of population susceptibility, as the assumptions underlying the range of plausible generative models resulting in the observed distribution of age of onset of FND. We combined these model families into 24 parametric proportional hazards models, and fitted each to the observed distribution of reported age at onset in two large FND datasets, one for functional movement disorders (FMD) and one for functional seizures (FS). Out-of-sample predictive accuracy for these models was compared using Bayesian model comparison.

**Results:** Strong trends were seen across model families with different distributions of population susceptibility to FND. For both datasets, the best-fitting model family overall was the mixture-cure family, which represents susceptibility as binary, with a susceptible and an unsusceptible proportion of the population. For the FMD dataset, some models in the log-normal frailty family had comparable fits to the mixture-cure models, and for the FS dataset, a number of the gamma frailty family had comparable fits. The variance parameters for each of these frailty distributions were so large as to imply binary risk, approximating mixture-cure models.

Models with exponential hazard distributions—which correspond to a generative process where a single trigger in a susceptible person brings about the condition—were universally poor fits for the observed data. Other hazard distributions were insufficiently distinguished by their out-of-sample predictive accuracy to make further inference as to the underlying process resulting in onset of FND in susceptible individuals.

**Interpretation:** Our results suggest that susceptibility to FND is approximately binary, with the susceptible proportion of the population extremely likely to develop FND in their lifetime. The results also argue strongly against a generative model where a single trigger is sufficient to cause the onset of FND in a susceptible person.

## Introduction

Understanding of the aetiology of functional neurological disorder (FND) has advanced markedly over the last two decades, with mainstream thinking moving from psychoanalytic conjecture to theories and empirical work invoking ideas and methodologies on the leading edge of neuroscience. Despite this, fundamental questions about the pathophysiology of the condition remain unanswered. Two such questions concern the population distribution of susceptibility to FND, and the sequence of events required to develop the condition. In other words, who can get FND, and what does it take for them to develop it?

Little is known about the distribution of underlying susceptibility to developing FND. Risk factors for the condition are known, some of which are acquired, such as adverse childhood experiences, depression, and anxiety;^1–3^ and some of which are neurodevelopmental, such as autism.^4,5^ The existence of identifiable risk factors suggests, at minimum, that susceptibility to FND is not homogeneous, but beyond this it is unclear whether susceptibility to FND is painted in shades of grey, or black and white.

The fact that FND is an acquired condition implies that at least one event needs to occur for the condition to manifest. Both physical insults and adverse life events have been shown to be extremely common in the days before onset of FND,^6,7^ and both have been suggested to be causative. Whether one or both of these is necessary to develop the condition, and whether a triggering event is sufficient, or merely the last in a required chain of events, remains unknown.

Gaining insight as to the population distribution of susceptibility to FND and the number of events necessary to develop the condition has the potential to focus, and therefore accelerate, research into the pathophysiology of the condition. Knowing the number of events required to develop FND could immediately eliminate pathophysiological theories which imply different numbers or natures of steps to what are expected. The distribution of population risk implies different approaches to both exploratory and hypothesis testing research on FND. If susceptibility is strongly inhomogeneous, or even binary, then three-way comparisons between people with FND, unaffected people who are susceptible to FND, and people unsusceptible to FND are likely to give more easily interpretable results for understanding the processes that cause symptoms and susceptibility than comparisons to a single control group. If, on the other hand, susceptibility is relatively homogeneous, in vivo research into the pre-symptomatic events that lead to FND (assuming such a process is necessary) will be difficult, and is likely to require innovative approaches; on the other hand, comparisons between people with FND and controls can more safely be assumed to reflect only differences related to the pathophysiology, and not to underlying susceptibility.

Here we identified six plausible mechanisms for the development of FND (Figure *1*). First, a single event might trigger development of the condition, and that event might occur with (1) constant, (2) monotonically changing, or (3) rising then falling probability across age. Second, a defined number of events might need to occur for the condition to develop, which might (4) arise as the result of parallel processes, or (5) have to occur sequentially. Finally, the condition might develop when (6) a variable which can rise and fall (for example, physiological stress) crosses a threshold.

**Figure 1:**
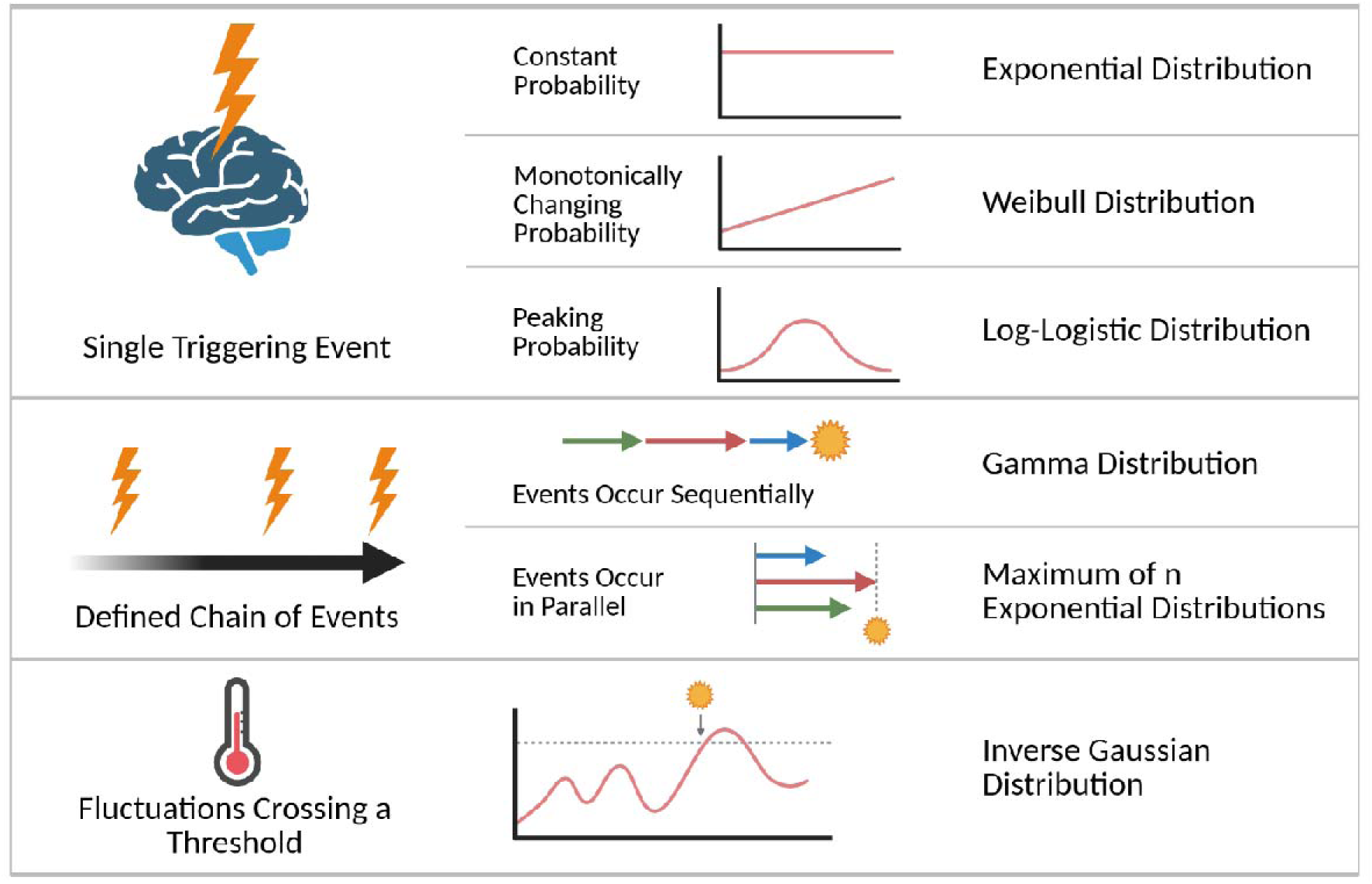
Plausible mechanisms of development of FND and their implied waiting time distributions for age of onset.

For susceptibility, we considered the possibilities to lie along a continuum from homogeneous risk across the population, through increasingly skewed levels of risk, to binary risk, where a subset of the population are able to develop FND and the remainder are not.

These fundamental questions of pathophysiology are not accessible at the individual level with our current understanding of FND. We therefore sought to use population-level data to address both questions, expanding a methodology which has been successful in other fields.^8,9^ Each possible process for developing FND suggests a distinct distribution for the age-specific incidence of the condition, based on the waiting time distribution family which corresponds to the generative process (Box 1). We constructed models corresponding to each combination of the susceptibility and hazard distributions outlined above, and fitted each to two published datasets on age of onset for FND. We then used Bayesian model comparison to compare the fits as evidence for the underlying generative models.

### Box 1: Survival Analysis and Generative Processes

#### Waiting Time Distributions

The expected waiting time for an event to occur is a function of the stochastic processes required to cause the event. Different causative processes will generate different waiting time distributions, and with sufficient data, these distributions can be used to make inferences about the underlying generative processes.

As an example, we can imagine a radiation detector which alarms when radiation is detected. We don’t know how the detector works, but based on our knowledge of other radiation detectors, we have two leading theories: (1) the detector alarms every time it detects a radioactive particle, or (2) the detector alarms after detecting a certain number () of radioactive particles. If we introduce a radioactive source (which emits particles with probability per unit time), and then repeatedly reset the detector and record the time until it alarms, each theory predicts a different distribution of waiting times. For the case where the detector alarms for each particle, the expected waiting time distribution is the exponential distribution, and for the case where multiple events are required, the expected waiting time is described by a gamma distribution with shape parameter. The figure below illustrates the difference in shape between these distribution families. By comparing observed data to these generative models, we can make inferences as to which generative model is more likely to be the cause of the data.

**Box 1.**
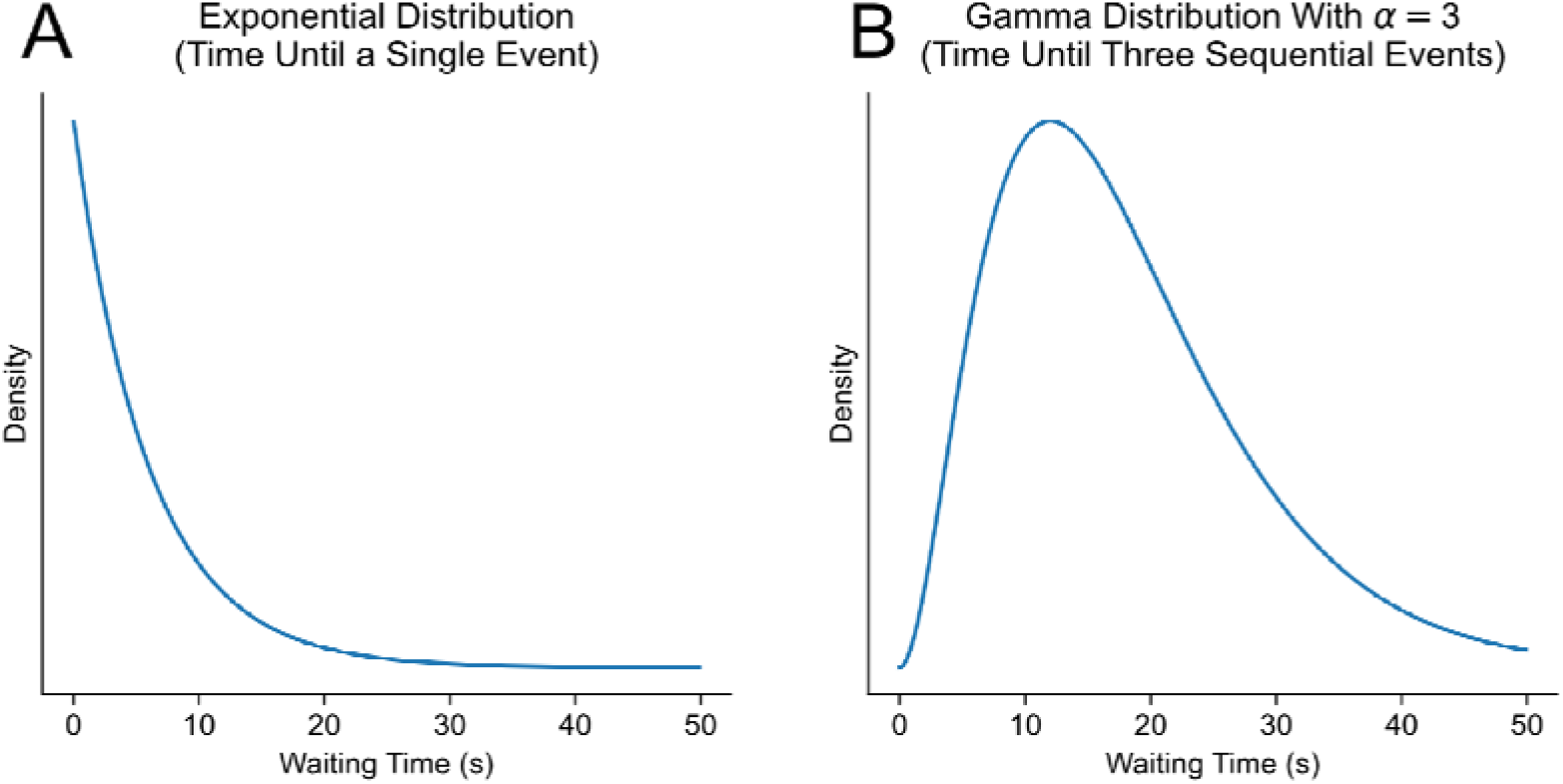
figure: Expected waiting times for events caused by (A) a one-step generative process, and (B) a generative process with three sequential events.

#### Population Susceptibility

Survival analysis models for populations assume both a hazard (waiting time) distribution, for the event of interest, and a distribution of risk (susceptibility) to that event across the population. In proportional hazards models, as used in this study, an individual’s risk for developing the condition at time is the population level hazard multiplied by their own individual susceptibility (drawn from the population distribution). The simplest possible population susceptibility distribution assumes equal susceptibility for all individuals. The situation where a proportion (IT) of individuals are equally susceptible to the condition and the remainder of the population are unable to develop it is described by mixture-cure models. Between these two extremes, frailty models assume a distribution of susceptibilities across the population. Here we used two different frailty models, one with population susceptibility described by a gamma distribution (used for its shape and analytical properties, not as a waiting time distribution as described above), and another using a log-normal distribution for population susceptibility. The log-normal distribution has a fatter tail, allowing a greater proportion of the population to have extreme values of susceptibility compared to the majority.

## Methods

### Model Formulation

For each of the possible mechanisms of FND development outlined above, we identified the waiting time distribution implied by the generative process (Figure *1*). The exponential distribution was used for a single event with constant probability; the Weibull distribution was used for a single event with monotonically changing probability; the log-logistic distribution was used for a single event with rising then falling probability; the gamma distribution was used for a defined number of events which must occur sequentially; the maximum of n exponential distributions, defined by the cumulative density function (CDF) of the exponential distribution to the power of n was used for a defined number of events which can occur independently; and the inverse Gaussian distribution was used for the time for a stochastically varying quantity to cross a threshold (the first passage time).

To capture the spectrum of possible distributions of susceptibility across the population, we considered four possibilities: uniform susceptibility across the population; a thin-tailed distribution of susceptibility across the population described by the Gamma distribution; a heavy-tailed distribution of susceptibility described by the log-normal distribution; and a situation of binary susceptibility, where a proportion of the population have uniform susceptibility to FND and the remainder are unable to develop it, described by a mixture-cure model.

To build models of age-specific incidence under each combination of possibilities, we constructed 24 parametric proportional hazards frailty models by combining each of the six hazard distributions describing disease development with one of the four population susceptibility families. Frailty distributions were parameterised with a fixed mean of 1 and variance as a free parameter. A Poisson likelihood distribution was used, with the rate parameter set for each bin as the remaining population at risk times the interval probability of developing FND (one minus the difference in value of the survival function at the bin edges).

Waiting time distribution parameters, frailty variance, and the cure fraction for the mixture-cure model were treated as free parameters. Prior means for these variables were set by simulating data from the model to find parameter values which gave a peak incidence in the first half of life, thin tails at the extremes of age, and estimated lifetime prevalence within an order of 2 of the observed value. Weakly informative dispersion parameters were used for priors. Analysis was performed using Python, with models specified and fitted using PyMC.^10^

### Data Preparation

Published distributions for age of onset by two-year bin for 4905 people with functional movement disorders^11^ (FMD) and 698 people with functional seizures^12^ (FS) were used to construct distributions of age-specific incidence for each subgroup. The available data for FMD were binned with a width of two years. For consistency, and to reduce noise, the FS dataset (which was in one-year bins) was converted to the same two-year width bins.

To construct age-specific incidence distributions for FND from these data, we needed an estimate of lifetime prevalence of the condition. This has been less studied than the incidence of the condition,^13^ so in addition to considering published estimates, we used actuarial tables and the distributions of ages of onset of FMD (the larger of the two datasets) to construct a meta-analytic estimate for lifetime prevalence based on annual incidence of the condition as estimated in the systematic review by Finkelstein et al.^13^ This gave a range which was concordant with published estimates of the lifetime prevalence, and which we believe is likely to be more accurate given the studies included.

To construct the dataset of age-specific incidence for use in our model, we used the mean estimated lifetime prevalence along with the US actuarial life table for the median year of birth of people in the FMD cohort to infer the implied total number of surviving people in each two-year bin.

As a sensitivity analysis assessing the impact of inaccuracy in our estimate of lifetime prevalence (and hence absolute age-specific incidence), we used the same methodology to construct datasets for the minimum and maximum estimates for annual incidence presented in Finkelstein et al^13^ and compared model-fits for these datasets to model-fits for our main FMD dataset.

### Model Comparison

We fitted each of our 24 models to the age-specific incidence datasets for both FMD and FS using Markov chain Monte Carlo sampling (MCMC). Convergence and sampling adequacy were assessed using standard diagnostics, with acceptable fits defined by R̂ ≤1.0, and sufficient effective sample sizes (bulk ESS > 200, and tail ESS > 100), consistent with current recommendations for Bayesian model evaluation.^14^

Out of sample predictive accuracy was assessed using leave one out cross validation (LOO-CV) with Pareto smoothed importance sampling. For datapoints where LOO might be unreliable (Pareto k > 0.7),^15^ exact LOO was performed by refitting the model with the corresponding observation omitted. Model comparison was based on the resulting expected log predictive density (ELPD).^16^

### Data Availability

The study used publicly available datasets. Analysis code is available through the Open Science Framework at https://doi.org/10.17605/OSF.IO/E4TDM.

## Results

### Overall Model Fits

Models varied widely in how well they represented the observed age-specific incidence and lifetime-prevalence. In-sample predictive fit is summarised in Figure 2 and Figure 3, which present the posterior predictive estimates for each model compared to the true incidence curves for FMD and FS. Out-of-sample predictive accuracy is summarised by the ELPD, which is presented per model for the FMD and FS datasets in Figure 4 and Figure 5.

**Figure 2:**
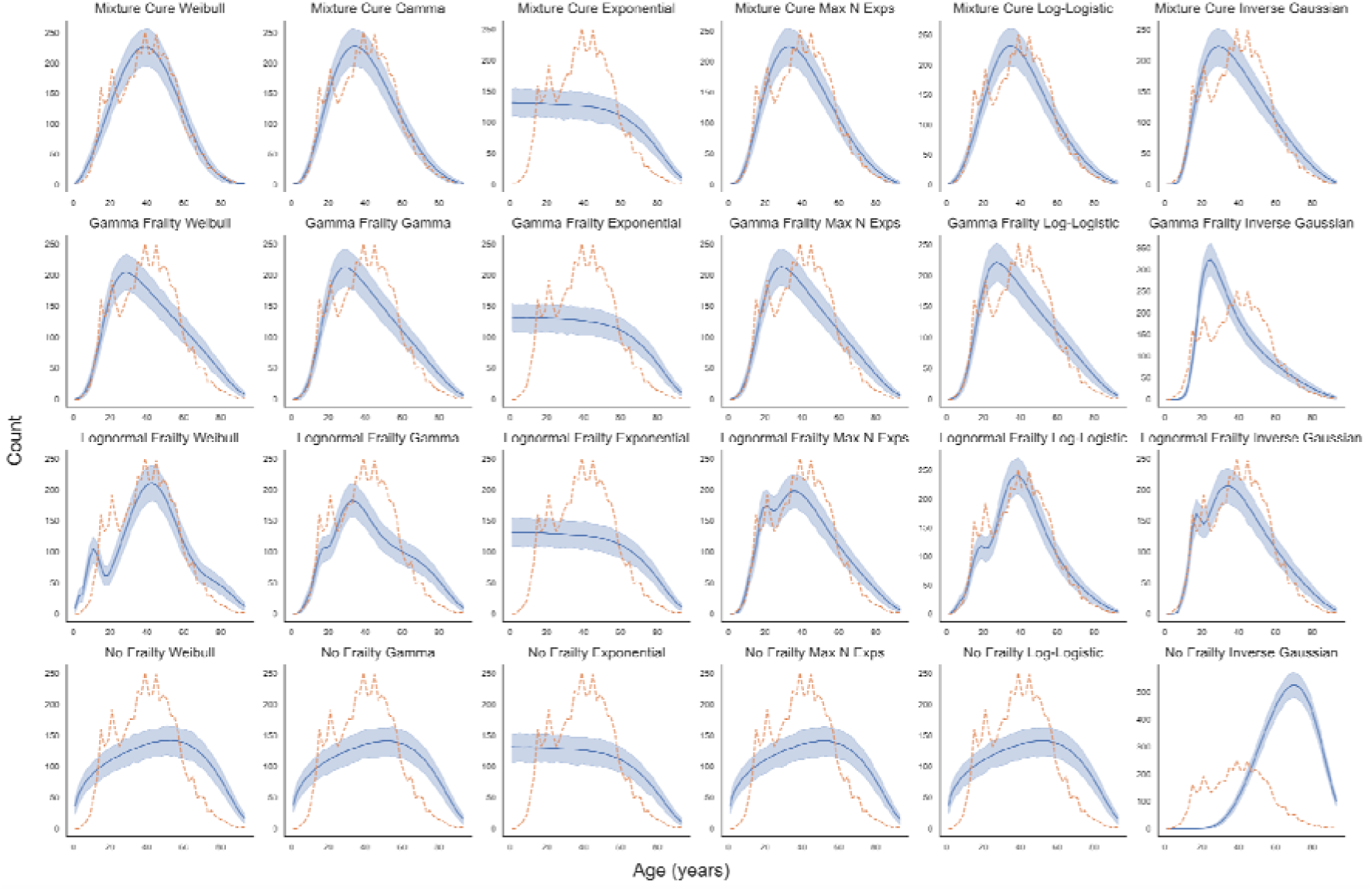
Posterior predictive estimates of age-specific incidence of FMD for each model, arranged by row for susceptibility distribution and by column for hazard distribution. The posterior mean for predicted incidence is plotted in blue, with shading representing 95% highest density interval. Observed incidence is plotted in orange.

**Figure 3:**
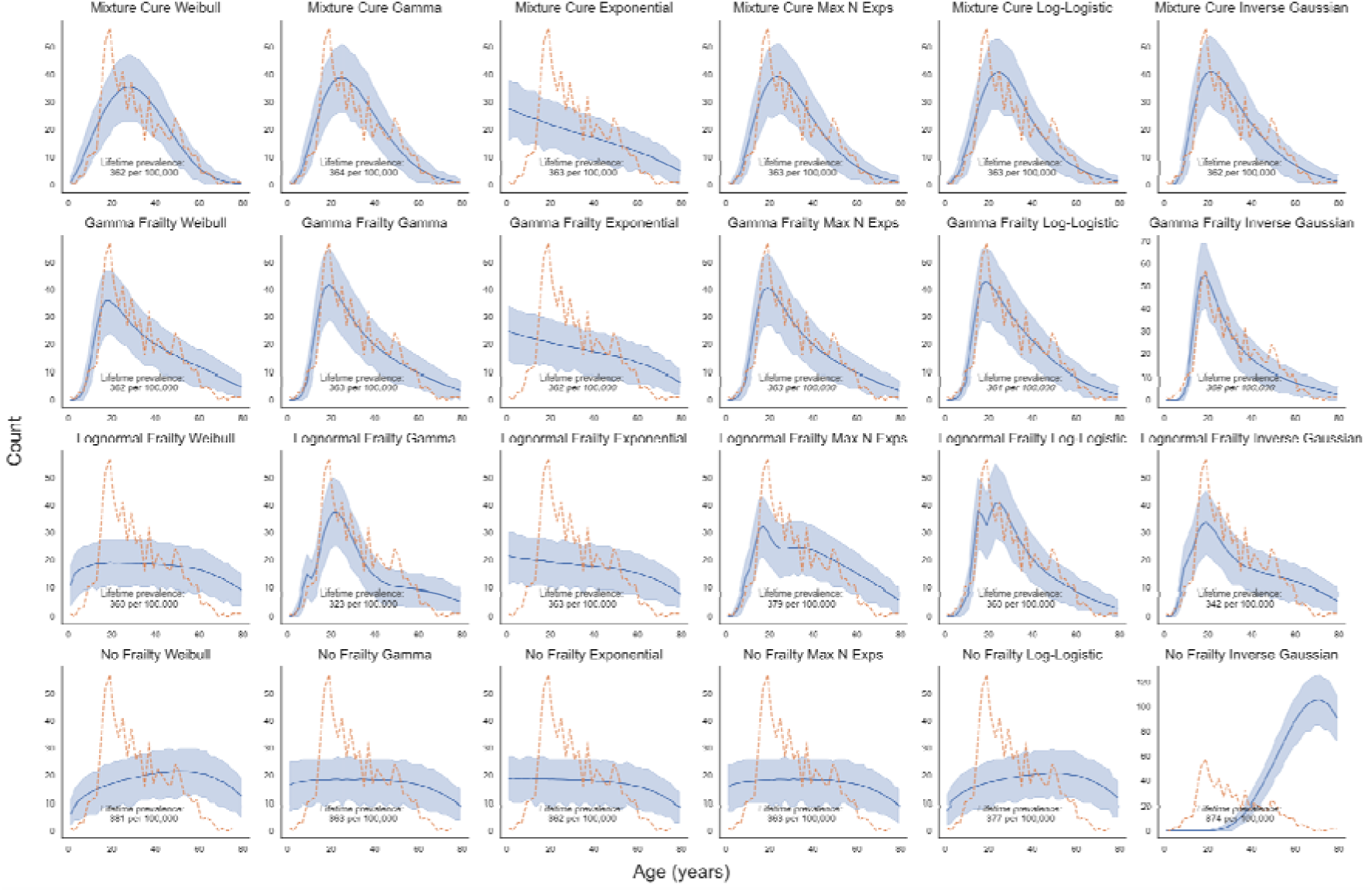
Posterior predictive estimates of age-specific incidence of FS for each model, arranged by row for susceptibility distribution and by column for hazard distribution. The posterior mean for predicted incidence is plotted in blue, with shading representing 95% highest density interval. Observed incidence is plotted in orange.

**Figure 4:**
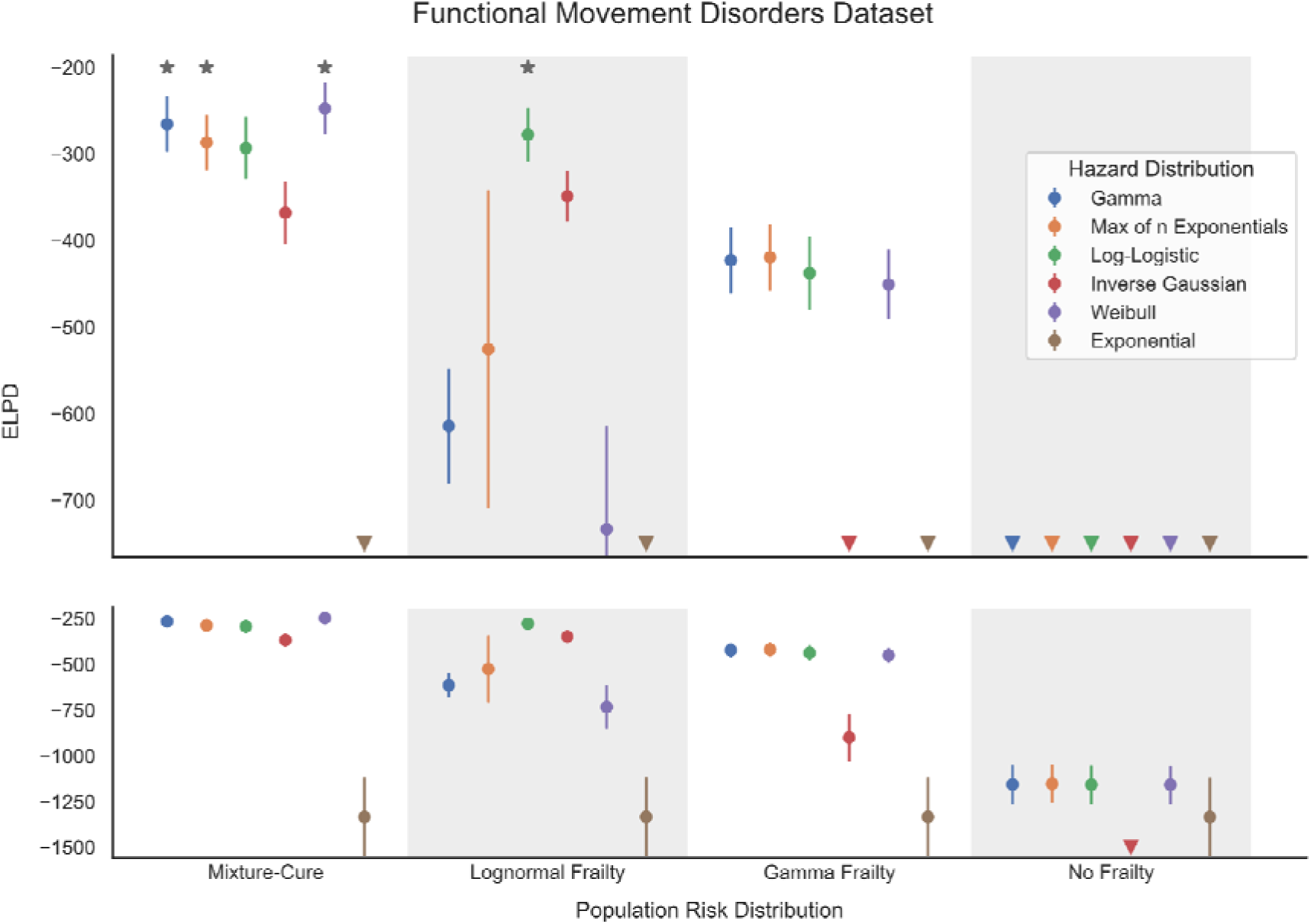
ELPD of each fitted model for the FMD dataset at two scales to allow visual inspection of relevant differences between models. Points represent mean ELPD with error bars indicating standard error. Triangles indicate that the value lies below the axis limit. Stars indicate models which are not formally distinguishable in fit from the best fitting model.

**Figure 5:**
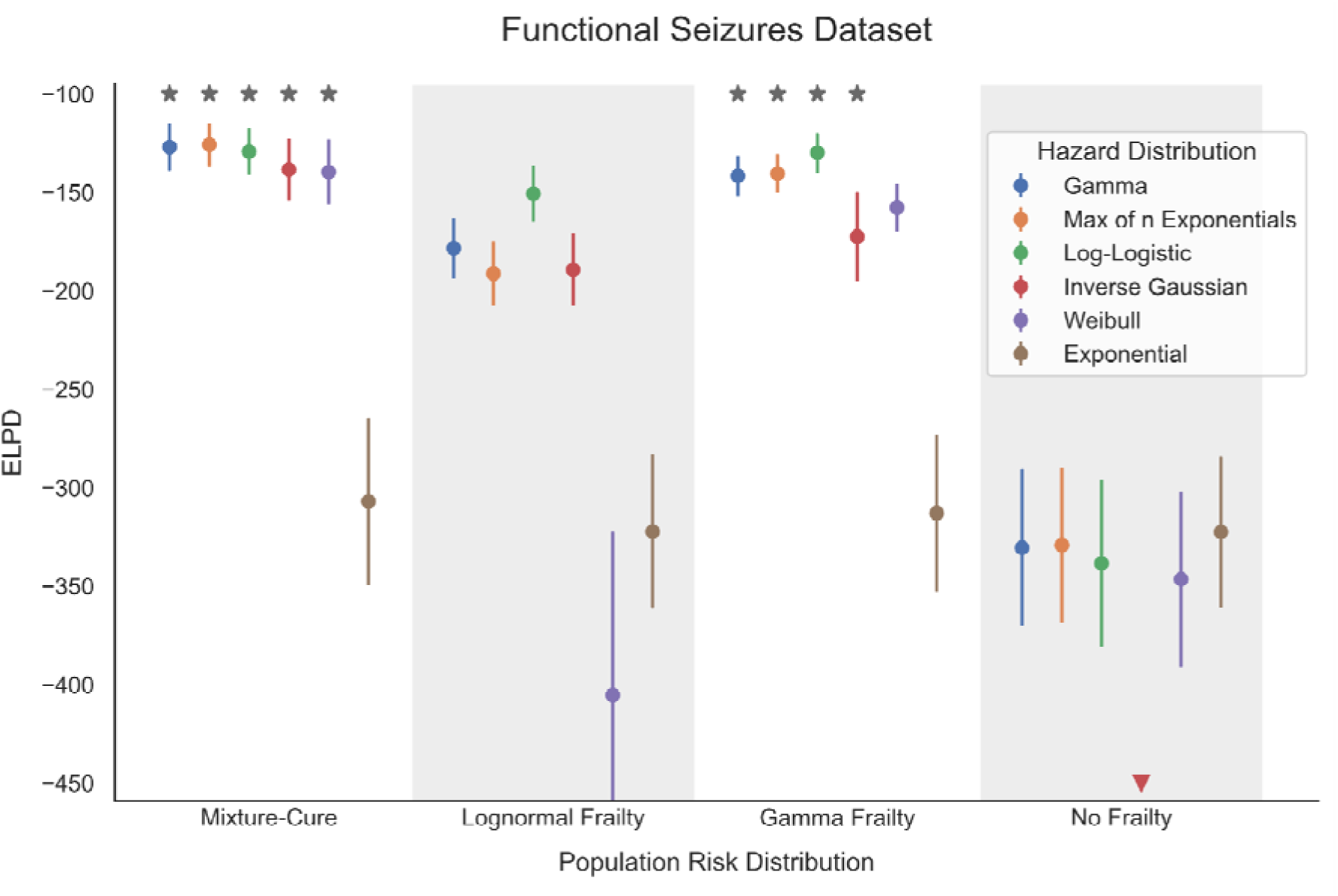
ELPD of each fitted model for the FS dataset. Points represent mean ELPD with error bars indicating standard error. Triangles indicate that the value lies below the axis limit. Stars indicate models which are not formally distinguishable in fit from the best fitting model.

Models which differed in ELPD by six or less, or by less than two times the standard error for difference (where the assumptions of the standard error calculation are met) were considered to be equivalent in out-of-sample predictive accuracy.^16^ By these criteria, four models jointly fitted the FMD data best: the mixture-cure models with Weibull, gamma, and maximum of exponential hazard distributions; and the log-normal frailty model with log-logistic hazard. For the FS dataset, nine models were indistinguishable in ELPD from the best fitting model. These were all five of the mixture-cure models excepting the exponential hazard model, and four of the six gamma frailty models: those with maximum of exponentials, gamma, log-logistic, and inverse Gaussian hazard distributions. Parameter estimates for the best-fitting distributions for the FMD dataset are provided in Supplementary Table 1, and for the FS dataset in Supplementary Table 2.

Some models were unable to meaningfully represent the observed data. Models with uniform population susceptibility, and models with exponential hazard distributions, were universally poor fits across both datasets, with large ELPD’s and minimal concordance with the shape of the age-specific incidence curve. These models should be considered excluded from general statements about model families made below.

Model families with mixture-cure population susceptibility were generally the best fits for the observed data, with all five (except the exponential hazard model) among the seven best fitting models for the FMD dataset, and all five among the models indistinguishable as best fitting for the FS dataset.

The FMD and FS datasets differ qualitatively in shape in early adulthood. Incidence in the FS dataset increases sharply in the late teenage years, then declines monotonically for the rest of adulthood. By contrast, the FMD dataset has a more gradual (although still abrupt) rise in incidence from childhood to early adulthood, and is bimodal, with a local maximum between ages 20 and 30, with a decline in incidence before increasing again to a global maximum at approximately age 40. Models of the lognormal frailty family were the only ones able to qualitatively capture the bimodality seen in the FMD dataset, although they did so with varying fidelity. The only model tied for best fit for the FMD dataset which did not use mixture-cure population susceptibility was the log-normal frailty log-logistic hazard model, and the log-normal frailty inverse Gaussian hazard model—which also qualitatively fitted this bimodality—was the next best fitting model. For the FS dataset, which did not have the bimodality seen in the FMD data, the gamma frailty model families were better fits overall than the log-normal frailty models, with four (excepting the Weibull and exponential hazards) among the nine models indistinguishable by ELPD.

Across both datasets, the fitted frailty variance parameters for the models of both frailty families (log-normal and gamma) implied extreme variation in susceptibility to FND across the population. The 95^th^ percentile of susceptibility was between two and five orders of magnitude greater than the median risk for the log-normal frailty models of both datasets, and tens to hundreds of orders of magnitude for the thinner tailed gamma-frailty models. With such large variances, the fits of these models could better be understood as approximating binary risk (as in the mixture-cure models) rather than the continua of risks across the population that they were intended to model.

For all mixture cure models except those with exponential hazard, the fitted parameter values for the unsusceptible proportion (IT) were within 0.18% of the complement of the observed lifetime prevalence for both datasets. This implies that, under these models, almost all individuals susceptible to FND will develop the condition in their lifetime.

Apart from the poor fit of the exponential models, family-level trends among models with different hazard distributions were much less pronounced. Across population susceptibility families in both datasets, the inverse Gaussian hazard family tended to be among the worst fits for the observed data, and showed extremely poor fits more frequently than other families. However, most differences were small compared with the differences seen between population susceptibility families.

### Sensitivity Analysis for Lifetime-Prevalence

To test the effect on our models of inaccuracy in the lifetime prevalence estimate used to generate population data, we constructed datasets for age-specific incidence of FMD using both the maximum and minimum estimates for lifetime prevalence, and fitted all models to each dataset. Supplementary figure 1 presents the ELPDs for each model for each dataset.

The lifetime-prevalence of the dataset used had minimal effect on ELPD for mixture-cure models, and no-frailty models, and for gamma frailty models, mildly affected only the fit of the poorly fitting inverse Gaussian hazard model, although this did not affect the model’s ranking compared to others.

Log-normal frailty models were somewhat more affected by the lifetime prevalence used, although only minor changes in overall model ranking resulted from this. In the log-normal frailty family, the maximum of n exponentials, inverse Gaussian, and Weibull hazard models had worse ELPD for both the minimum and maximum lifetime prevalence datasets; the log-logistic hazard model had worse ELPD for the maximum lifetime prevalence dataset, but no meaningful change with the minimum lifetime prevalence dataset; and the Gamma frailty model had worse ELPD with the minimum lifetime prevalence dataset, and improved fit with the maximum lifetime prevalence dataset.

## Discussion

We constructed a set of models corresponding to the set of commonly discussed or assumed generative models of FND with respect to the distribution of susceptibility and the processes culminating in its development,^17–20^ and compared them to observed data from large datasets of age at onset of the condition. In doing so, we were able to show that these models vary markedly in probability with respect to the observed data. The results provide evidence that can be used to support some of these commonly assumed models, and to discount others.

Taken together, the results suggest that susceptibility to developing FND is distributed across the population in an approximately binary fashion which is best approximated by mixture-cure susceptibility models. Furthermore, the fitted parameter values for our mixture-cure models suggest that the majority of the population who are susceptible to FND will develop it within their lifetimes.

The fits of the models we specified were unable to distinguish between most hazard distributions, but they did provide strong evidence against the exponential distribution as a description for the generative process underlying FND. Importantly, this distribution, which describes the expected time to occurrence of a single random event that occurs with fixed probability over time, is often tacitly assumed in the discussion of FND, since it represents the case where the condition is caused directly by triggers such as injuries, illnesses or stress.^17–20^ The failure of this model to describe the data suggests that the occurrence of one of these triggers in a susceptible person is unlikely to be sufficient as a causative process, and that these triggers (presuming they are mechanistically involved) must interact with other processes to cause the onset of the condition. We have argued elsewhere that FND might be better conceived of as a failure of the mechanisms which resolve the transient, mild functional symptoms that occur in healthy people than as the process which generates the symptoms themselves.^21^ If that is the case, then triggering events might frequently result in functional symptoms, but only lead to FND when they occur in concert with other processes which prevent resolution of the symptoms.

Both datasets we used had distinctive features in the shape of their age at onset curves, which were difficult for models to fit. The FS dataset has a sharp increase in incidence between the mid- and late-teenage years, with relatively low incidence in childhood. In contrast, the FMD dataset has a more gradual increase in incidence from childhood to adulthood, but is bimodal, with a peak in incidence between the ages of 14 and 22 years, with a decline before rising again to the true modal age of onset between 38 and 40 years.

The FS dataset comes from the recorded ages of onset of symptoms for participants in a large treatment study for FS.^12^ The trial only included participants aged 18 years and over who still had functional seizures. Since remission rates are high in children with FND^22^ only a minority of people with childhood-onset FND would be expected to have had their symptoms persist long enough to be included in the study. It is therefore likely that the FS dataset underestimates the incidence of FND in childhood, which leads to the apparent abrupt increase in incidence just below the age limit for inclusion.

The FMD dataset was generated by meta-analysis of both published studies and unpublished data from many sites over many countries, which would be expected to reduce systematic bias. The reason for the bimodality in the dataset is unclear. Increased incidence in this period would not be inconsistent with our clinical experience, and may represent the true distribution of ages of onset. Events around the affected ages could also plausibly cause artefact such as this though. The handover of care from paediatric to adult neurologists during the affected period might affect the reported age-at-onset in a number of ways, and it is worth noting that the study predominantly included data obtained from studies of adults and from adult neurologists. Salient events which mark time in those years (school years, leaving home, university, etc.) might also plausibly serve as mnemonic anchors, causing recall bias in people with longstanding symptoms. This would be expected to cause an artefactual increase in reported incidence at these ages, with relative decreases in surrounding age categories, which visually appears possible after this spike. On balance, we believe there is equipoise as to whether this bimodality is a property of the true distribution or is artefactual, but that it is more likely than not to be genuine.

In summary, our results suggest that susceptibility to FND across the population is extremely heterogeneous, approximating binary risk, with a small subgroup susceptible to and very likely to develop FND, while the majority of the population are extremely unlikely to develop the condition. It also provides evidence against models where a single trigger leads to development of the condition in a susceptible person. Further use of this methodology with larger, higher quality datasets that may be available in the future might allow for better distinction between hazard distributions and their implied generative models of disease development.

