## Supplementary material for "Who Can Get Functional Neurological Disorder and How Do They Get It? Pathophysiological Insights from Epidemiological Data"

| Susceptibility distribution  Hazard distribution | Parameter | Mean (95% HDI) |
| --- | --- | --- |
| Mixture-Cure |  |  |
| Gamma | $\pi$ (cure fraction) | 0.9935 (0.9933—0.9937) |
|  | $\mu$ (hazard mean) | 44.9 (44.1—45.7) |
|  | $\alpha$ (shape) | 4.44 (4.24—4.65) |
| Maximum of n exponentials | $\pi$ (cure fraction) | 0.9932 (0.9930—0.9934) |
|  | $\lambda$ (scale) | 20.8 (20.0—21.7) |
|  | $\alpha$ (number of events) | 5.0 (4.7—5.3) |
| Weibull | $\pi$ (cure fraction) | 0.9937 (0.9935—0.9939) |
|  | $\lambda$ (scale) | 47.8 (47.1—48.4) |
|  | $k$ (shape) | 2.60 (2.54—2.67) |
| Log-normal frailty |  |  |
| Log-logistic | $\sigma$ (frailty shape) | 5.80 (5.76—5.83) |
|  | $\alpha$ (scale) | 40.5 (39.2—41.7) |
|  | $\beta$ (shape) | 4.7 (4.5—4.8) |

Table 1: Distribution parameter estimates for the best fitting models for the FMD dataset.

| Susceptibility distribution  Hazard distribution | Parameter | Mean (95% HDI) |
| --- | --- | --- |
| Mixture-Cure |  |  |
| Gamma | $\pi$ (cure fraction) | 0.9962 (0.9959—0.9965) |
|  | $\mu$ (hazard mean) | 32.7 (31.4—34.1) |
|  | $\alpha$ (shape) | 4.21 (3.75—4.70) |
| Maximum of n exponentials | $\pi$ (cure fraction) | 0.9961 (0.9958—0.9964) |
|  | $\lambda$ (scale) | 5.01 (4.33—5.76) |
|  | $\alpha$ (number of events) | 5.0 (4.3—5.8) |
| Log-logistic | $\pi$ (cure fraction) | 0.9960 (0.9957—0.9963) |
|  | $\alpha$ (scale) | 30.4 (29.0—31.8) |
|  | $\beta$ (shape) | 3.09 (2.85—3.32) |
| Inverse Gaussian | $\pi$ (cure fraction) | 0.9960 (0.9958—0.9963) |
|  | $\mu$ (mean) | 35.0 (32.8—37.4) |
|  | $\lambda$ (shape) | 103 (91—118) |
| Weibull | $\pi$ (cure fraction) | 0.9962 (0.9959—0.9965) |
|  | $\lambda$ (scale) | 36.3 (35.1—37.7) |
|  | $k$ (shape) | 2.29 (2.15—2.43) |
| Gamma frailty |  |  |
| Gamma | $\theta$ (frailty variance) | 5.5e-4 (4.9e-4—6.2e-4) |
|  | $\mu$ (hazard mean) | 88 (73—106) |
|  | $\alpha$ (shape) | 6.3 (5.2—7.2) |
| Maximum of n exponentials | $\theta$ (frailty variance) | 6.0e-4 (5.3e-4—6.7e-4) |
|  | $\lambda$ (scale) | 46 (36—58) |
|  | $\alpha$ (number of events) | 5.9 (4.9—6.8) |
| Log-logistic | $\theta$ (frailty variance) | 4.4e-4 (4.0e-4—4.8e-4) |
|  | $\alpha$ (scale) | 52.1 (47.5—56.8) |
|  | $\beta$ (shape) | 6.0 (5.55—6.50) |
| Inverse Gaussian | $\theta$ (frailty variance) | 7.3e-4 (6.6e-4—8.0e-4) |
|  | $\mu$ (mean) | 1558 (525—2630) |
|  | $\lambda$ (shape) | 188 (180—196) |

Table 2: Distribution parameter estimates for the best fitting models for the FMD dataset.

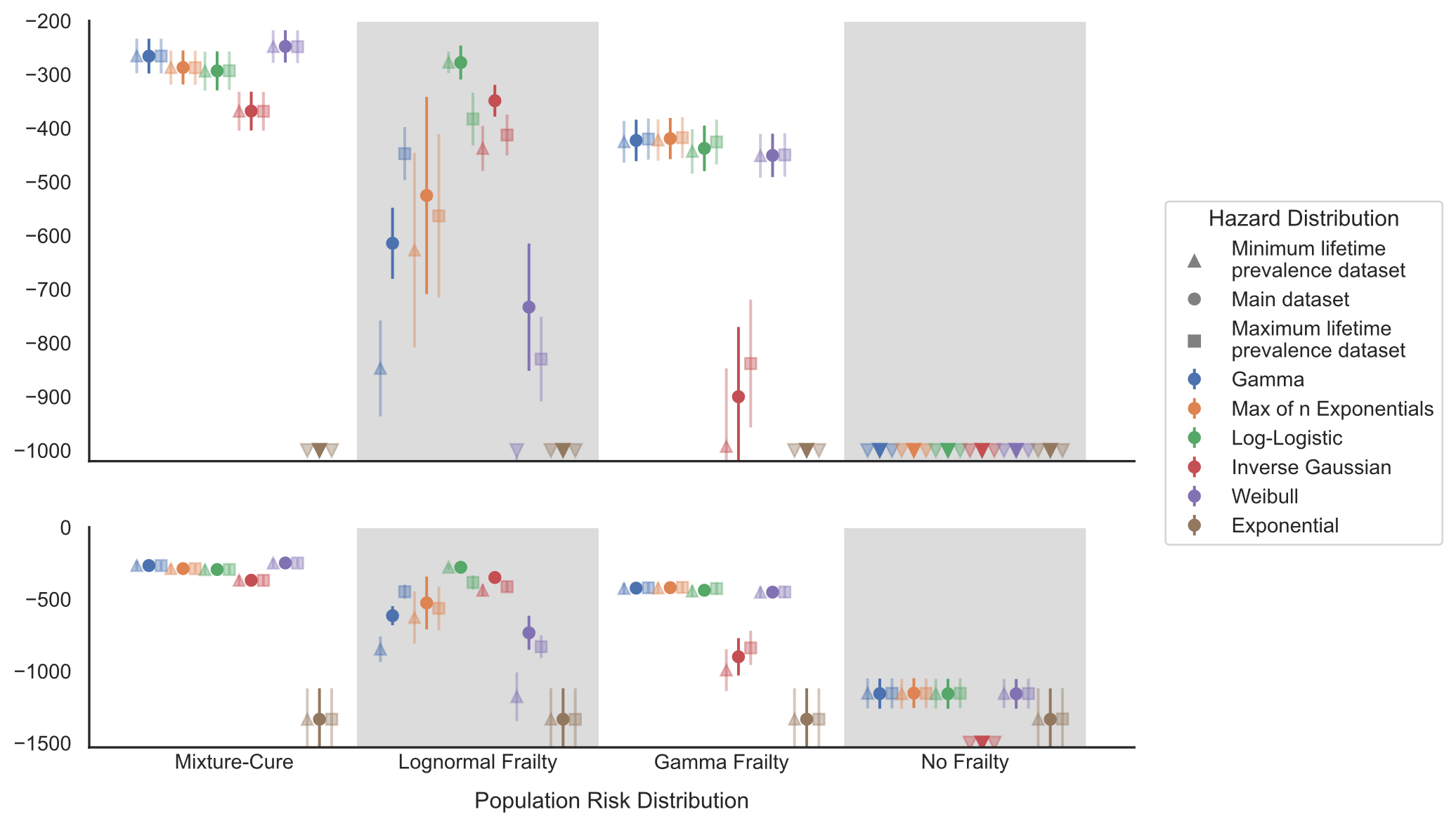

Supplementary Figure 1: Sensitivity analysis showing ELPDs of models when fitted to FMD datasets constructed using the minimum (apex-up triangles), mean (circles), and maximum (squares) estimates for lifetime prevalence. Points represent mean ELPD and error bars represent standard error. Plots present the same data at two scales to allow relevant visual comparisons.
